# Differential Associations of Interleukin 6 Receptor Variant Across Genetic Ancestries and Implications for Targeted Therapies

**DOI:** 10.1101/2022.09.24.22280325

**Authors:** Xuan Wang, Isabelle-Emmanuella Nogues, Molei Liu, Tony Chen, Xin Xiong, Clara-Lea Bonzel, Harrison Zhang, Chuan Hong, Kumar Dahal, Lauren Costa, J. Michael Gaziano, Seoyoung C. Kim, Yuk-Lam Ho, Kelly Cho, Tianxi Cai, Katherine P. Liao

**Affiliations:** Department of Biostatistics, Harvard T.H. Chan School of Public Health, Boston, MA 02115, USA; VA Boston Healthcare System, Boston, MA 02130, USA; Harvard Medical School, Boston, MA 02115; Brigham and Women’s Hospital, Boston, MA 02115, USA; Duke University, Durham, NC, USA

## Abstract

Genomic data are increasingly incorporated into high-throughput approaches such as the Phenome-Wide Association Study (PheWAS) to query potential effects of targeted therapies. Genetic variants, such as the interleukin-6 receptor (*IL6R*) genetic variant rs2228145 (Asp358Ala), have been identified with a downstream effect similar to the drug, e.g., tocilizumab which targets IL6R, and can be used to screen for potential protective or harmful signal across a broad range of traits in large biobanks with linked genomic and clinical data. To date, there are limited approaches to determine whether these effects may differ across diverse populations to inform potential differential drug effects especially in populations under-represented in clinical trials. In this study, we developed and applied an approach to detect heterogeneous associations, using the *IL6R* variant as an example, in African vs European ancestry. We identified a total of 29 traits with a differential association between the *IL6R* variant, with notable differences including a lower risk of type 2 diabetes in AFR vs EUR, and a higher white blood cell count. With the increasing use of targeted blockade of the IL6 pathway in conditions ranging from rheumatologic to cardiovascular conditions, the findings from this study can inform ongoing studies targeting IL6; general approach to test for heterogeneity of associations can be applied broadly to any PheWAS.

## Introduction

The interleukin-6 receptor (*IL6R*) single nucleotide polymorphism (SNP) (rs2228145, Asp(358)Ala) is associated with several conditions including a reduced risk for cardiovascular disease (CVD), peripheral arterial disease, and an increased risk for rheumatoid arthritis (RA) [1, 2]. This missense variant is known to reduce expression of IL6R, thereby reducing IL-6 signaling and downstream inflammation [3]. Therapies such as tocilizumab block the IL6R pathway and is indicated for the treatment of inflammatory conditions such as RA and large vessel vasculitis and hospitalized COVID-19. In a phenotypic screen, subjects carrying the Asp(258)Ala variant were found to have a phenotypic profile similar to those on drugs that block IL-6R; subjects with the *IL6R* variant have higher hemoglobin and lower high sensitivity C-reactive protein (CRP) compared to those without the variant. In randomized clinical trials in RA, treatment with IL6R dramatically reduced hsCRP [4, 5]. Earlier studies also demonstrated IL-6 infusions in humans reduced hemoglobin counts while cessation enabled a return to baseline [6]. In a phenotypic screen for *IL6R*, similar associations were observed whereby individuals carrying the *IL6R* variant had lower hsCRP and higher hemoglobin. This proof-of-concept study suggested that studying phenotypes associated with *IL6R* may also inform associations that may not be detected from clinical trials based on predetermined endpoints. With increasing interest in targeting the IL6 pathway in the general population for reduction of CV risk, continued use in COVID-19, and ongoing studies in RA [7, 8], large datasets containing linked genomic and phenotype data can provide clues for considering potential differences in effect across populations. Moreover, there is a growing need for methods that can distill information from high dimensional data to provide information on heterogeneity based on effect size and significance.

The objective of this study was to develop and apply an approach to systematically identify heterogeneous associations in African vs European ancestry, the two largest ancestries in a diverse mega-biobank cohort, as part of an *IL6R* PheWAS. We hypothesize that this large-scale screen will identify differential effects of the *IL6R* variant across phenotypes with implications for current and future trials targeting the IL6 pathway. Findings were validated in two independent biobank cohorts.

## Methods

### Study Populations

We performed an *IL6R* PheWAS in the Veterans Affairs Million Veteran Program (MVP) cohort with data up to 09/30/2020 and findings were replicated in UK Biobank (UKB) and the Mass General Brigham (MGB) Biobank [9, 10, 11].

The VA MVP is a longitudinal, multi-institutional cohort study that collects clinical Electronic Health Records (EHR) data, namely inpatient and outpatient data combined with genomic data from participants in approximately 50 Veterans Affairs facilities across the United States. Subjects were included in the MVP if they were 18 years of age or older; had a valid mailing address (to ensure the possibility of follow-up); were able to provide informed consent at the time of recruitment. All participants were required to provide written informed consent upon recruitment. They were asked to 1) complete baseline and lifestyle questionnaires, providing information such as self-reported race/ethnicity, dietary habits, and smoking/drinking status, as well as 2) provide blood samples for genotyping and biomarker studies. This study obtained institutional review board approval through the Veterans Affairs MVP.

The UKB is a longitudinal cohort study that prospectively recruits patients to determine the effects of lifestyle, environmental, and genomic factors on disease outcomes over time. The study population includes approximately 500,000 volunteers recruited from the United Kingdom’s general population from 2006 to 2010. Measurements of 61 laboratory biomarkers and blood cell counts were ascertained for all UKB participants as part of a standardized baseline assessment.

The MGB Biobank contains linked EHR, and genetic data anchored by two large tertiary care hospitals: Brigham and Women’s Hospital and Massachusetts General Hospital in Boston. The MGB Biobank data consist of 59,052 participants with both EHR data and genomic data available. Laboratory test results were extracted for these patients.

### Statistical Methods

The PheWAS analysis was performed using a standardized approach [12]. Briefly, we fitted a logistic regression for PheWAS analysis to test for association with phenotypes as defined by PheCodes and linear regression for the laboratory analysis. Since many of the laboratory measurements are highly skewed, we tested for association of the *IL6R* variant with log-transformed laboratory values. All models were adjusted for patient age, sex, length of EHR follow up, and health care utilization as measured by the log-total number of PheCodes.

Genetic ancestry was ascertained using commonly used methods. Briefly, we trained a logistic regression classification algorithm using self-reported race as silver standard labels and 127 ancestry informative SNPs [13]. The cut-off of predicted probabilities for classification is chosen to guarantee sensitivity is above 0.975. We excluded related MVP participants (halfway between second-degree and third-degree relatives or closer) as measured by the Kinship-Based Inference for GWAS software. We stratified all association analyses of the *IL6R* variant, rs2228145 (risk allele A; Asp358Ala), with disease phenotypes and laboratory test results by the predicted ancestry group. We focused the analyses on the two largest ancestry groups in MVP, African (AFR) and European (EUR) ancestry.

Within each ancestry group, we performed PheWAS analyses including 1,875 phenotypes as defined by PheCodes [14] and 69 routine laboratory measurements curated in prior studies at the VA, which includes complete blood count and lipid profiles. For each phenotype, a participant was defined as having the condition if they had at least 2 PheWAS codes. We excluded PheWAS codes with a prevalence of 0.5% or less from the analysis and excluded integer level (parent) PheCodes for which corresponding descendant PheCodes already existed, leaving a total of 660 remaining phenotypes. For example, we excluded the integer PheCode 250 (Diabetes mellitus) but included the descendant PheCodes such as 250.1 (Type 1 Diabetes) and 250.2 (Type 2 Diabetes). The screen was also performed on 69 adjudicated laboratory measurements available at VA. Values were defined by the median of all available measurements for each patient. A detailed list of the laboratory tests is in the Supplementary Materials Table S5. We first compared associations between *IL6R* and PheCodes and separately for the curate laboratory values in AFR vs EUR. Significant PheCodes/labs within each ancestry were determined with a false discovery rate (FDR) <0.1 using the Benjamini-Hochberg procedure [15].

Heterogeneity testing was conducted to identify phenotypes and laboratory values to detect a differential association between *IL6R* and phenotype among AFR vs EUR ancestries. To adjust for multiple testing, we developed a novel false discovery rate (FDR) controlled heterogeneity testing (hetFDR) procedure which leverages information from both the mean effect and the magnitude of heterogeneity under a prior assumption that heterogeneous effects are more likely to be present for phenotypes with non-zero mean effects across a large number of candidate phenotypes. The hetFDR procedure is a three-step procedure. In Step (I), for each phenotype, we construct (i) an overall mean effect test statistic as an inverse-variance weighted average effect estimate combining the regression coefficients (against the genetic variant of interest) from the two ancestry groups along with its associated P value; as well as (ii) a chi-square test statistic ascertaining the heterogeneity between the effects as observed from the regression coefficients of the two groups. The mean effect statistic and the heterogeneity statistic are designed to be asymptotically independent so that the validity of tests is ensured when incorporating the mean effect statistics to assist the heterogeneity testing. In Step (II), we use the mean effect statistics to weight the heterogeneity P values, assigning higher prior probabilities of null hypothesis rejection to those phenotypes with more significant mean effects, which corresponds to our prior assumption that phenotypes with non-zero mean effects are more likely to show heterogeneity across the considered ancestry groups. The weighting function is decided adaptively from the data through a regression-based approach. In the final Step (III), we adopt the multiple testing procedure of [16] on the weighted heterogeneity p-values for detection with FDR control. We controlled for an FDR of 10%, which ensures that among the associations considered significant, at most 10% of the associations were false positives [15]. A detailed description for the statistical method of heterogeneity testing is provided in the Supplementary Materials: Statistical Methodology.

### Replication of laboratory results using UK Biobank and MGB Biobank Data

To validate heterogeneous *IL6R*-phenotype associations in AFR vs EUR observed in MVP, we performed analyses in UKB and MGB Biobank data. Due to the relatively smaller size of AFR in these cohorts, the analyses focused on traits with continuous values, i.e., laboratory results.

This study was approved by the institutional review boards of the VA Boston Healthcare System and Mass General Brigham. All analyses were performed using R software.

## Results

A total of 545,147 Veterans were included in the analysis, of which 91.3% were men, with a mean (SD) age of 62.1 (13.9) years and a mean (SD) follow-up time of 12.5 (5.7) years. Among these participants, 105,838 were classified as AFR and 439,309 were classified as EUR.

Overall, we observed 10 phenotypes with significant associations with *IL6R* among Veterans of AFR ancestry compared to 34 among Veterans of EUR ancestry, none of which were significant in both populations (Figure 1). For laboratory measurements, we observed 30 measurements with significant associations with *IL6R* among Veterans of AFR ancestry compared to 28 among Veterans of EU ancestry (Figure 2). *IL6R* was significantly associated with 18 labs across both ancestries.

**Figure 1.**
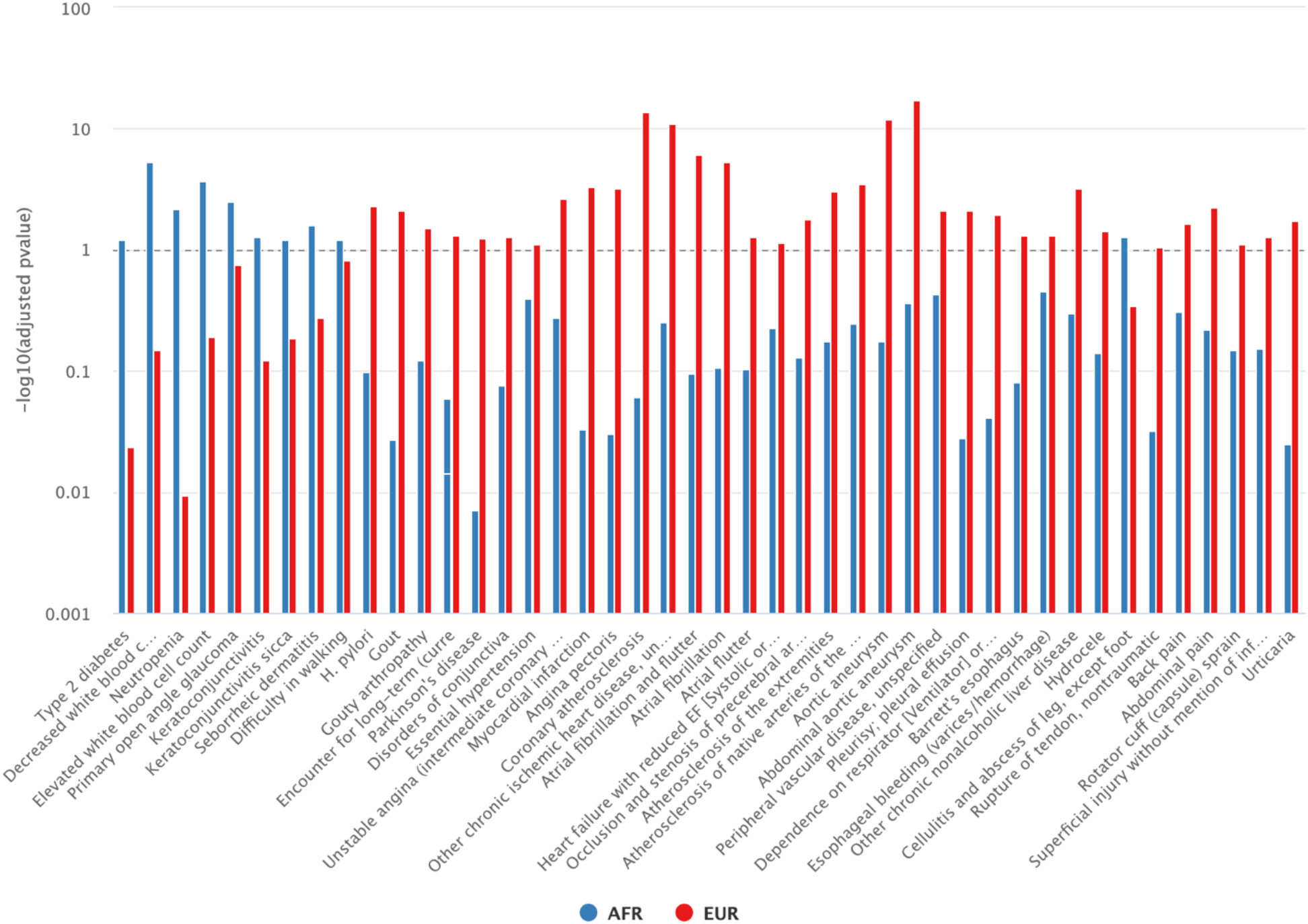
Phenotypes (phecodes) significantly associated with the *IL6R* variant in AFR or EUR (BH adjusted p value <= 0.1)

**Figure 2.**
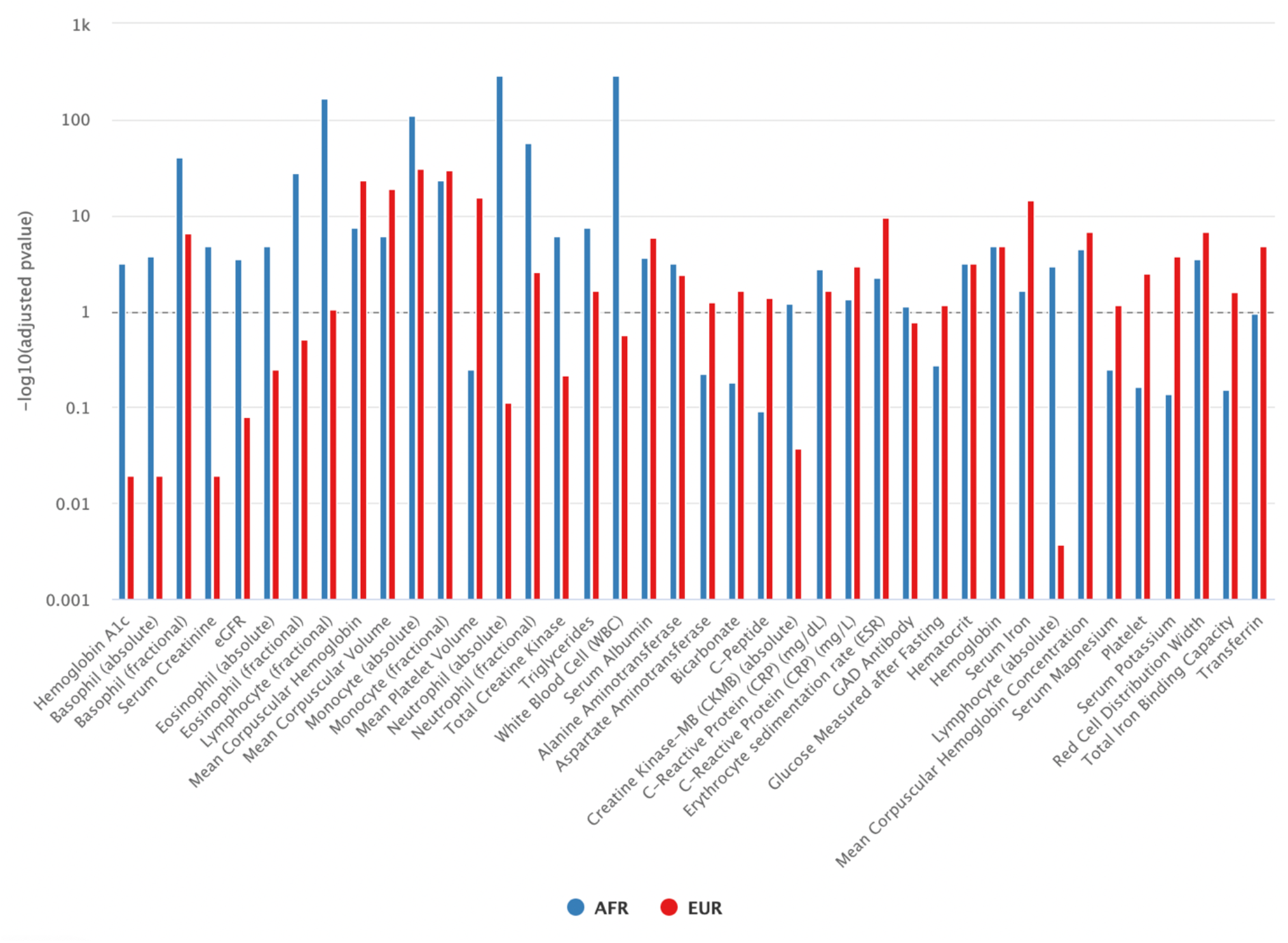
Laboratory measurements significantly associated with the *IL6R* variant in AFR or EUR (BH adjusted p value <= 0.1)

The majority of *IL6R*-phenotype associations within the EUR pertained to vascular and cardiac disease. The phenotypes with the strongest association with *IL6R* were aortic aneurysm (odds ratio [OR], 0.92; 95% CI, 0.90-0.94) as well as a specific type of aortic aneurysm, abdominal aortic aneurysm (AAA) (OR, 0.89; 95% CI, 0.87-0.90), coronary atherosclerosis and ischemic heart disease (OR, 0.96; 95% CI, 0.95-0.97) (Figure 1 and 2). The corresponding associations in AF are similar but not significant [(AA) OR=0.95 (0.87-1.03); (AAA) OR=0.89 (0.80-1.00); (CHD) OR=0.99 (0.95-1.02)]

After applying the test for heterogeneity, we observed 11 PheCodes translating to 7 conditions with differential association in AFR vs EUR: glaucoma, keratoconjunctivitis, periodontitis, type 2 diabetes, seborrheic dermatitis, walking difficulties, white blood cell count elevation (Figure 3). *IL6R* was associated with reduced odds for glaucoma, keratoconjunctivitis, periodontitis, and type 2 diabetes among AFR with either no association or increased odds in EUR. The *IL6R* variant was associated with higher odds of an elevated white blood cell count in AFR (OR1.21, 95% CI 1.12-1.30), and in line with this, a lower odds ratio for neutropenia in AFR (OR 0.80, 95% CI 0.72-0.89); these associations were not observed among EUR. *IL6R* was associated with seborrheic dermatitis and difficulty walking with increased odds in AFR and reduced odds in EUR.

**Figure 3.**
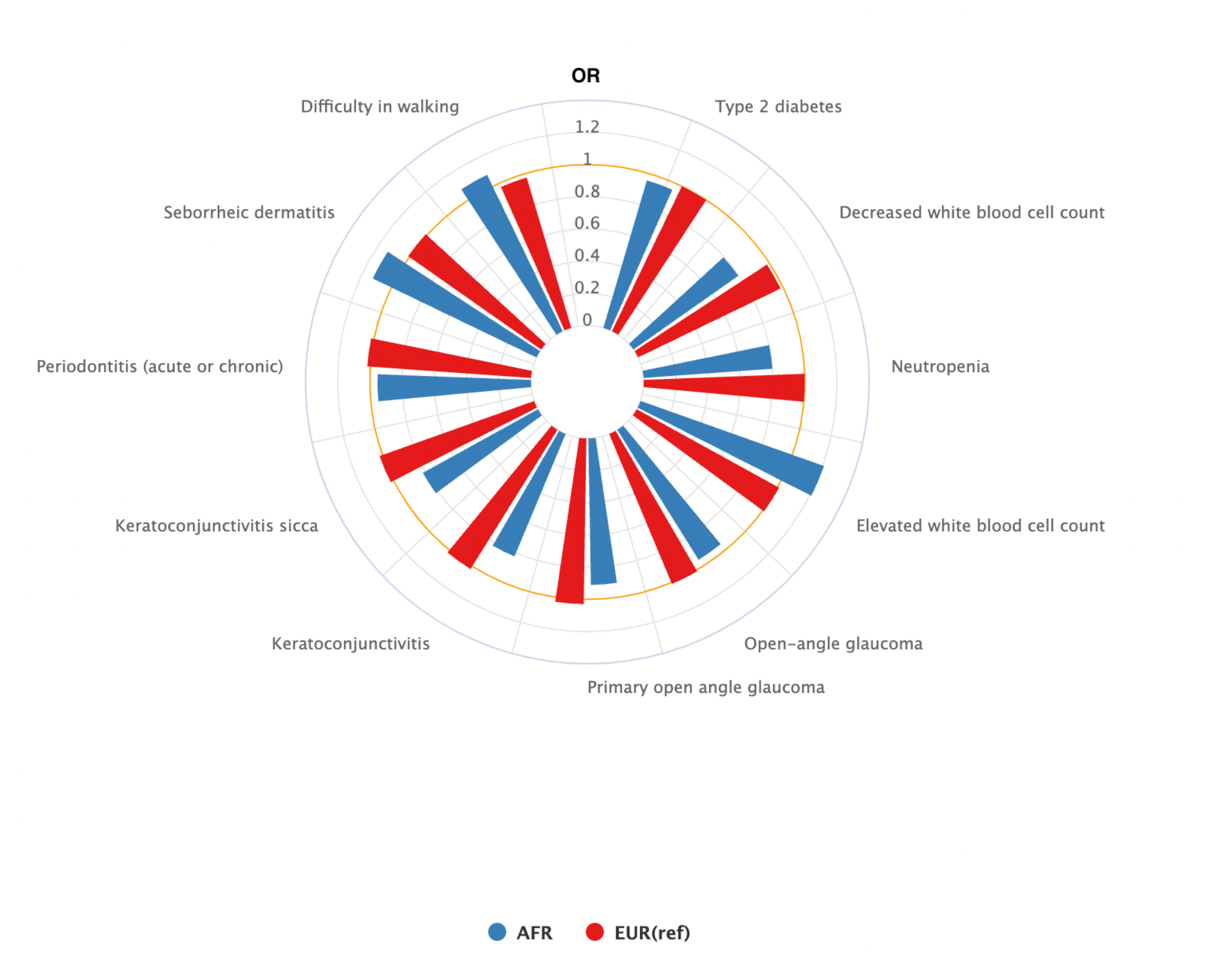
Odds ratios for phenotypes with significant differential associations in AFR vs EUR ancestries (BH adjusted p value <= 0.1)

A comparison of laboratory values identified differences across 18 laboratory measurements (Figure 4). In line with the significant difference in ICD codes related to WBC, the largest difference was observed in WBC whereby among individuals of AFR ancestry, each copy of the *IL6R* variant was associated with a higher WBC compared to those who did not carry the variant; no association was observed between *IL6R* and WBC among EUR. The higher value was observed across neutrophils, monocytes, eosinophils, and basophils, with the difference was most pronounced in absolute neutrophil count; the *IL6R* variant was associated with higher absolute values of neutrophils in AFR vs EUR. *IL6R* was also associated with higher triglyceride levels in AFR compared to EUR. The variant was associated with lower hemoglobin a1c (hba1c) in AFR with no significant association observed in EUR, in line with a lower odds ratio of T2D observed in AFR.

**Figure 4.**
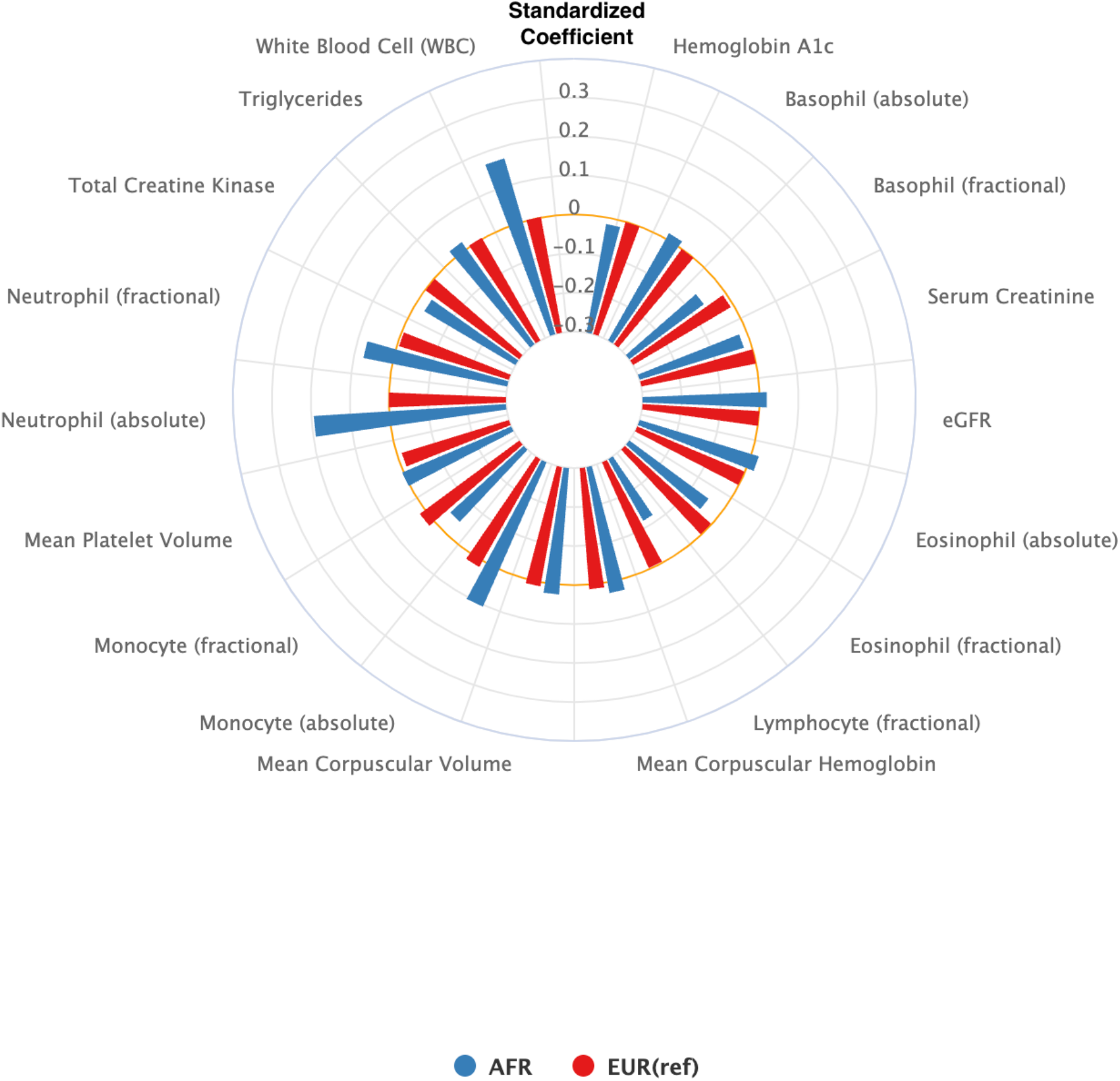
Comparison of standardized coefficients for associations between *IL6R* with laboratory values in AFR vs EUR (BH adjusted p value <= 0.1)

Due to the limited cohort size of individuals of AFR ancestry in the UKB and MGB, validation was focused on replicating laboratory values.

The association and differences in WBC in AFR vs EUR remained the most significant finding. *IL6R* was associated with higher WBC among individuals of AFR vs EUR in both cohorts (Supplementary Table S3 and S4). *IL6R* was also associated with higher triglycerides in AFR vs EUR across the replication cohorts.

Due to strong and differential associations between *IL6R* with white blood cell phenotypes, we further tested the association between the variant and serious infection stratified by ancestry [17]. Overall, we observed an association between *IL6R* and a modest but significantly increased odds of serious infection in AFR but not EUR [AFR OR 1.03, 95% CI 1.01-1.04 vs EUR with OR 1.01, 95% CI 1.00-1.01]. Due to the small population size in UKB and MGB we did not have sufficient power to validate in these populations.

## Discussion

This study provides a new roadmap for leveraging large biobanks to screen for differential associations genetic variants and phenotypes across a diverse population. These data in turn can be used to inform potential differential effects of targeted therapies using an application designed to test for heterogeneity in large-scale genotype-phenotype screens. We focused on a specific variant in *IL6R* with the known downstream effect of reducing IL-6 signaling with effects similar to existing therapies targeting IL-6. In a prior study, our group demonstrated the proof of concept that the associations identified from a PheWAS of the *IL6R* variant mirror clinical and laboratory findings from clinical trials of treatment with IL6R. As an example, treatment with IL6R antagonists is known to significantly reduce hsCRP levels; individuals carrying the IL6R variant have lower hsCRP levels than those who do not. In this study using the most recent data from MVP, a biobank with the largest population of individuals of AFR ancestry in the world, we observed 29 traits with heterogeneous associations, including WBC and T2D.

The most significant heterogeneous signal observed was a lower odds ratio of neutropenia or higher WBC among Veterans of AFR descent compared to EUR; in EUR no association was observed between *IL6R* and WBC. The clinical significance of the association between *IL6R* and higher WBC, particularly neutrophil counts in AFR and EUR ancestry is unclear. To provide context, in a large population-based epidemiologic study, WBC was lower in Black compared White individuals [18]. As WBC are involved in host defense, in the present study, we tested the association between *IL6R* and serious infection and observed a modest but significant increased odds for serious infection among individuals of AFR descent where no association was observed in EUR. We were underpowered to validate these findings in UKB or MGB. In a review of the literature, we were unable to identify clinical trials of therapies targeting IL6 studies stratifying outcomes or adverse events by self-reported race (as genetic ancestry data are typically not available in trials). The majority of large observational studies for infection risk and IL6R blockade stems from studies of tocilizumab, the first IL6R antagonist approved for use in the US for RA. In these studies, risk of infection on tocilizumab is compared with another targeted therapy and overall, no difference has been observed [19, 20], however there were no data stratifying by self-reported race or ethnicity. Based on findings from the present study, we anticipate that in studies with adequately sized populations, we would anticipate higher WBC among individuals of AFR ancestry on IL6R blockade, as well as a small increased odds for serious infection. Future trials and studies on the IL6 pathway can consider collecting data on WBC and neutrophil count, as well as stratifying infectious adverse events by self-reported race.

The heterogeneity test also identified an association between the *IL6R* variant with a reduced odds of T2D among Veterans of AFR descent, while no association was observed in EUR. Likewise, hba1c which reflects an average level of glucose over 2-3 months, was lower among individuals of AFR carrying the *IL6R* variant, while no association was observed among EUR in MVP. A lower hba1c was also observed among AFR carrying the *IL6R* variant compared to EUR in UKB. To our knowledge, glucose and hba1c levels were not reported in the randomized controlled trials in rheumatoid arthritis or giant cell arteritis [21, 22, 23]. However, the general association between the *IL6R* variant and lower odds of T2D was observed in meta-analysis examining the potential role of this pathway in the etiology of T2D [24, 25]. Additionally, higher serum IL6 levels are associated with higher levels of hba1c, and increased risk of developing T2D in a large cohort study of women [25, 26]. In an observational cohort study of RA patients with hba1c measurements before and after initiation of tocilizumab compared to a tumor necrosis factor inhibitor, a larger reduction in hba1c was observed in the tocilizumab group [27]. Thus, our study corroborates these findings and further anticipates that individuals of AFR descent either with T2D or at risk of T2D may derive more benefit from IL6R compared to individuals of EUR descent.

Notably, the strong associations observed between the *IL6R* variant and cardiovascular phenotypes, e.g. coronary heart disease, aortic aneurysms, peripheral arterial disease observed in prior studies was confirmed in EUR but not AFR [28, 29, 30]. This difference in association between *IL6R* and cardiovascular phenotypes in AFR vs EUR did not reach statistical difference with regards to heterogeneity. The hetFDR approach leverages information from both the mean effect and the magnitude of heterogeneity to determine the significance of the differences based on data from the entire population. Thus, in comparison to other phenotypes studied, the differential association with CV phenotypes were not considered heterogeneous and we would not anticipate a significant difference in the salutary effect of IL6R blockade for CV phenotypes in AFR vs EUR.

The hetFDR procedure applied in this study for multiple testing of heterogeneity fills an unmet need for methods that allow us to screen high-throughput data efficiently, such as PheWAS for differences across diverse patient populations. Compared with existing commonly used FDR control approaches like Benjamini and Hochberg’s procedure (BHq) [15] and Stoery’s procedure [31], our method is more powerful in detecting the phenotypes with heterogeneous effects. HetFDR takes advantage of the fact that among all phenotypes, only a small fraction has non-zero effects and nearly all those phenotypes with heterogeneous effect tend to have non-zero mean effects on the whole population, which can be characterized more effectively compared to the heterogeneity due to the larger sample size. This property was confirmed with our simulation results given in the Supplementary Materials. Specifically, we demonstrated in a simulation study using a similar scale of data and variable types as our current biobank datasets, the hetFDR achieved a satisfactory FDR control and a uniformly higher power compared to other existing methods.

### Limitation

The population sizes for individuals of AFR ancestry were significantly lower in the UKB and MGB biobanks compared to MVP (UKB, AFR: n = 7,538; EUR: n = 459,315; MGB, AFR: n=2,922; EUR: n=49,883; MVP, AFR: n = 105,838; EUR: n = 439,309). The smaller population resulted in limited power to replicate binary phenotypes, e.g., phecodes. Another potential limitation or difference between UKB and MVP is that UKB primarily contains inpatient codes and data from general medicine practices with less capture from other outpatient specialty practices in comparison to MVP and MGB. Importantly, this study did not include individuals of other ancestries.

## Conclusion

In summary, we leveraged 3 large population-based biobanks and applied a novel approach to test for heterogeneity identifying differential associations of the *IL6R* variant in AFR vs. EUR ancestry. Since the effect of the *IL6R* variant on phenotypic traits is known to parallel the effects of existing therapies targeting IL6R, findings from this study can inform ongoing and future trials targeting this pathway in the general population, particularly CVD. Our results suggest that targeting IL6R may be associated with higher WBC count and a potential modest signal for higher infection risk among individuals of AFR vs EUR descent. IL6R blockade may have a more beneficial effect for T2D with lower hba1c levels in AFR vs EUR, as well as potential beneficial effects for glaucoma, keratoconjunctivitis, and periodontitis. Notably, we observed a paucity of clinical trial data that were either sufficiently powered or reported data enabling post-hoc analyses of potential differences in effect across race and ethnicity. The increasing data available from more diverse populations such as MVP, along with the advancements in methods to analyze these data, can provide guidance on data elements to collect for pre-planned clinical trial subgroup analyses. Ultimately, these data together with approaches such as hetFDR can help us to design efficient trials that are to study the effectiveness of not just the primary outcome, but also potential beneficial and detrimental effects of a given therapy across a diverse population.

## Data Availability

All data produced in the present study are available upon reasonable request to the authors

## Acknowledgements

This work was funded by the US Veterans’ Health Administration Million Veterans Program (MVP), the Harvard School of Public Health’s Department of Biostatistics, and the Harvard Medical School’s Department of Biomedical Informatics, and the NIH P30 AR072577.

## Supplementary Materials

**Table S1.** Significant heterogeneous associations between the IL6R variant and phecode based phenotypes in MVP in AFR vs EUR, reduced odds in light red, increased odds in blue.

|  | African |  |  | European |  |  |  |
| --- | --- | --- | --- | --- | --- | --- | --- |
| Variable | OR | 95% CI | pval | OR | 95% CI | pval | heter.pval |
| others |  |  |  |  |  |  |  |
| Infection count | 1.028 | (1.013, 1.042) | 1.596 × 10<sup>-4</sup> | 1.007 | (1.001, 1.012) | 1.662 × 10<sup>-2</sup> | 1.428 × 10<sup>-2</sup> |
| musculoskeletal |  |  |  |  |  |  |  |
| Difficulty in walking | 1.083 | (1.033, 1.136) | 9.195 × 10<sup>-4</sup> | 0.975 | (0.956, 0.994) | 1.142 × 10<sup>-2</sup> | 9.145 × 10<sup>-3</sup> |
| dermatologic |  |  |  |  |  |  |  |
| Seborrheic dermatitis | 1.132 | (1.061, 1.207) | 1.799 × 10<sup>-4</sup> | 1.012 | (0.994, 1.029) | 1.853 × 10<sup>-1</sup> | 6.948 × 10<sup>-2</sup> |
| digestive |  |  |  |  |  |  |  |
| Periodontitis (acute or chronic) | 0.953 | (0.919, 0.99) | 1.233 × 10<sup>-2</sup> | 1.021 | (1.004, 1.038) | 1.453 × 10<sup>-2</sup> | 6.948 × 10<sup>-2</sup> |
| sense organs |  |  |  |  |  |  |  |
| Open-angle glaucoma | 0.953 | (0.925, 0.982) | 1.774 × 10<sup>-3</sup> | 1.012 | (0.999, 1.026) | 6.609 × 10<sup>-2</sup> | 3.607 × 10<sup>-2</sup> |
| Primary open angle glaucoma | 0.911 | (0.874, 0.95) | 1.407 × 10<sup>-5</sup> | 1.026 | (1.005, 1.047) | 1.552 × 10<sup>-2</sup> | 3.186 × 10<sup>-4</sup> |
| Keratoconjunctivitis | 0.824 | (0.738, 0.919) | 5.497 × 10<sup>-4</sup> | 1.016 | (0.977, 1.056) | 4.231 × 10<sup>-1</sup> | 4.085 × 10<sup>-2</sup> |
| Keratoconjunctivitis sicca | 0.817 | (0.725, 0.921) | 9.072 × 10<sup>-4</sup> | 1.024 | (0.981, 1.07) | 2.823 × 10<sup>-1</sup> | 4.228 × 10<sup>-2</sup> |
| hematopoietic |  |  |  |  |  |  |  |
| Decreased white blood cell count | 0.796 | (0.736, 0.86) | 7.989 × 10<sup>-9</sup> | 0.983 | (0.95, 1.018) | 3.458 × 10<sup>-1</sup> | 3.186 × 10<sup>-4</sup> |
| Neutropenia | 0.802 | (0.722, 0.891) | 4.222 × 10<sup>-5</sup> | 1.002 | (0.959, 1.048) | 9.250 × 10<sup>-1</sup> | 1.811 × 10<sup>-2</sup> |
| Elevated white blood cell count | 1.207 | (1.121, 1.299) | 6.449 × 10<sup>-7</sup> | 1.015 | (0.99, 1.041) | 2.518 × 10<sup>-1</sup> | 3.056 × 10<sup>-3</sup> |
| endocrine/metabolic |  |  |  |  |  |  |  |
| Type 2 diabetes | 0.956 | (0.93, 0.982) | 9.091 × 10<sup>-4</sup> | 1.001 | (0.991, 1.011) | 8.369 × 10<sup>-1</sup> | 8.458 × 10<sup>-2</sup> |

**Table S2.** Significant heterogeneous associations between IL6R with median laboratory values in AFR vs EUR ancestry in MVP; comparisons with heterogeneous p-value< 0.1 were considered significant, negative association in light red, and positive association in blue.

| Variable | African |  | European |  | heter.pval |
| --- | --- | --- | --- | --- | --- |
|  | ES | pval | ES | pval |  |
| Basophil - absolute | 0.017 | $4.184 \times 10^{-5}$ | 0.000 | $9.343 \times 10^{-1}$ | $5.304 \times 10^{-2}$ |
| Basophil - fractional | -0.062 | $2.907 \times 10^{-42}$ | -0.009 | $5.163 \times 10^{-8}$ | $3.639 \times 10^{-27}$ |
| eGFR | 0.019 | $9.669 \times 10^{-5}$ | 0.000 | $6.972 \times 10^{-1}$ | $4.509 \times 10^{-2}$ |
| Eosinophil - absolute | 0.020 | $3.370 \times 10^{-6}$ | -0.001 | $4.232 \times 10^{-1}$ | $9.944 \times 10^{-4}$ |
| Eosinophil - fractional | -0.047 | $1.722 \times 10^{-29}$ | -0.002 | $1.744 \times 10^{-1}$ | $1.624 \times 10^{-22}$ |
| Hemoglobin A1c | -0.017 | $2.118 \times 10^{-4}$ | 0.000 | $9.420 \times 10^{-1}$ | $7.332 \times 10^{-2}$ |
| Lymphocyte - fractional | -0.118 | $4.277 \times 10^{-167}$ | -0.003 | $3.445 \times 10^{-2}$ | $1.301 \times 10^{-141}$ |
| Mean Corpuscular Hemoglobin | 0.029 | $3.995 \times 10^{-9}$ | 0.012 | $4.140 \times 10^{-25}$ | $2.690 \times 10^{-3}$ |
| Mean Corpuscular Volume | 0.026 | $1.101 \times 10^{-7}$ | 0.011 | $3.029 \times 10^{-21}$ | $1.748 \times 10^{-2}$ |
| Mean Platelet Volume | 0.004 | $3.609 \times 10^{-1}$ | -0.012 | $2.063 \times 10^{-17}$ | $7.171 \times 10^{-3}$ |
| Monocyte - absolute | 0.105 | $1.852 \times 10^{-113}$ | 0.018 | $3.827 \times 10^{-33}$ | $1.884 \times 10^{-71}$ |
| Monocyte - fractional | -0.045 | $2.291 \times 10^{-25}$ | 0.017 | $4.476 \times 10^{-32}$ | $1.721 \times 10^{-40}$ |
| Neutrophil - absolute | 0.194 | $3.557 \times 10^{-322}$ | 0.001 | $6.243 \times 10^{-1}$ | $6.009 \times 10^{-299}$ |
| Neutrophil - fractional | 0.076 | $2.245 \times 10^{-58}$ | -0.005 | $6.291 \times 10^{-4}$ | $1.410 \times 10^{-57}$ |
| Serum Creatinine | -0.020 | $4.120 \times 10^{-6}$ | 0.000 | $9.240 \times 10^{-1}$ | $2.690 \times 10^{-3}$ |
| Total Creatine Kinase | -0.028 | $1.032 \times 10^{-7}$ | -0.001 | $4.703 \times 10^{-1}$ | $4.517 \times 10^{-4}$ |
| Triglycerides | 0.022 | $3.105 \times 10^{-9}$ | 0.004 | $6.187 \times 10^{-3}$ | $7.495 \times 10^{-4}$ |
| White Blood Cell (WBC) | 0.170 | 0.000 | 0.002 | $1.337 \times 10^{-1}$ | $5.537 \times 10^{-301}$ |

**Table S3.** Validation results in the MGB Biobank for significant heterogeneous laboratory findings in AFR vs EUR in MVP.

| Variable | African |  |  | European |  |  |
| --- | --- | --- | --- | --- | --- | --- |
|  | ES | 95% CI | pval | ES | 95% CI | pval |
| Basophil - absolute | 0.013 | (-0.03, 0.057) | $5.457 \times 10^{-1}$ | 0.000 | (-0.011, 0.012) | $9.916 \times 10^{-1}$ |
| Basophil - fractional | 0.036 | (-0.02, 0.092) | $2.070 \times 10^{-1}$ | -0.008 | (-0.026, 0.009) | $3.293 \times 10^{-1}$ |
| eGFR | -0.010 | (-0.032, 0.013) | $4.044 \times 10^{-1}$ | 0.000 | (-0.003, 0.004) | $8.147 \times 10^{-1}$ |
| Eosinophil - absolute | 0.028 | (-0.047, 0.104) | $4.614 \times 10^{-1}$ | -0.005 | (-0.026, 0.016) | $6.501 \times 10^{-1}$ |
| Eosinophil - fractional | -0.043 | (-0.098, 0.012) | $1.270 \times 10^{-1}$ | -0.002 | (-0.016, 0.011) | $7.461 \times 10^{-1}$ |
| Hemoglobin A1c | -0.003 | (-0.014, 0.009) | $6.586 \times 10^{-1}$ | 0.000 | (-0.003, 0.002) | $6.932 \times 10^{-1}$ |
| Lymphocyte - fractional | -0.050 | (-0.076, -0.025) | $1.000 \times 10^{-4}$ | -0.004 | (-0.01, 0.003) | $2.458 \times 10^{-1}$ |
| Mean Corpuscular Hemoglobin | 0.006 | (0.001, 0.012) | $1.790 \times 10^{-2}$ | 0.001 | (0, 0.002) | $2.140 \times 10^{-2}$ |
| Mean Corpuscular Volume | 0.005 | (0, 0.009) | $3.080 \times 10^{-2}$ | 0.001 | (0, 0.001) | $3.640 \times 10^{-2}$ |
| Mean Platelet Volume | 0.002 | (-0.004, 0.008) | $5.310 \times 10^{-1}$ | -0.001 | (-0.002, 0) | $4.370 \times 10^{-2}$ |
| Monocyte - absolute | 0.042 | (0.011, 0.074) | $8.600 \times 10^{-3}$ | 0.007 | (-0.002, 0.015) | $1.136 \times 10^{-1}$ |
| Monocyte - fractional | -0.009 | (-0.035, 0.017) | $5.014 \times 10^{-1}$ | 0.008 | (0.001, 0.014) | $1.650 \times 10^{-2}$ |
| Neutrophil - absolute | 0.094 | (0.066, 0.122) | 0.000 | 0.006 | (0, 0.012) | $3.580 \times 10^{-2}$ |
| Neutrophil - fractional | -0.076 | (-0.187, 0.035) | $1.821 \times 10^{-1}$ | -0.017 | (-0.036, 0.002) | $8.550 \times 10^{-2}$ |
| Serum Creatinine | -0.081 | (-0.187, 0.026) | $1.377 \times 10^{-1}$ | -0.005 | (-0.02, 0.01) | $5.084 \times 10^{-1}$ |
| Total Creatine Kinase | -0.077 | (-0.157, 0.003) | $6.040 \times 10^{-2}$ | -0.016 | (-0.033, 0) | $5.060 \times 10^{-2}$ |
| Triglycerides | 0.034 | (0.003, 0.066) | $3.190 \times 10^{-2}$ | -0.002 | (-0.009, 0.005) | $6.025 \times 10^{-1}$ |
| White Blood Cell (WBC) | 0.058 | (0.039, 0.077) | 0.000 | 0.004 | (0, 0.008) | $3.570 \times 10^{-2}$ |

**Table S4.**
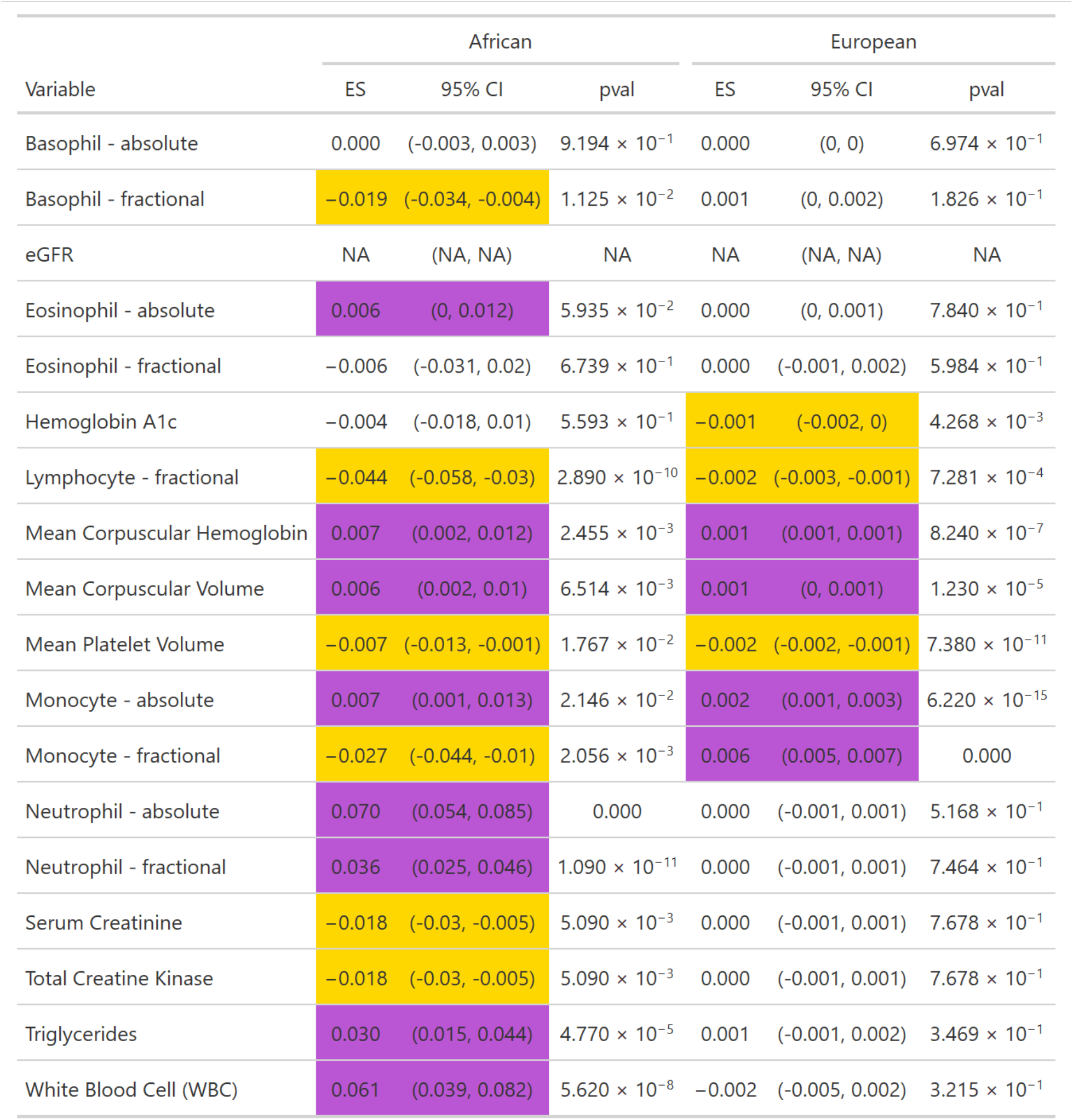
Validation results in UKB for significant heterogeneous laboratory findings in AFR vs EUR in MVP.

**Table S5.**
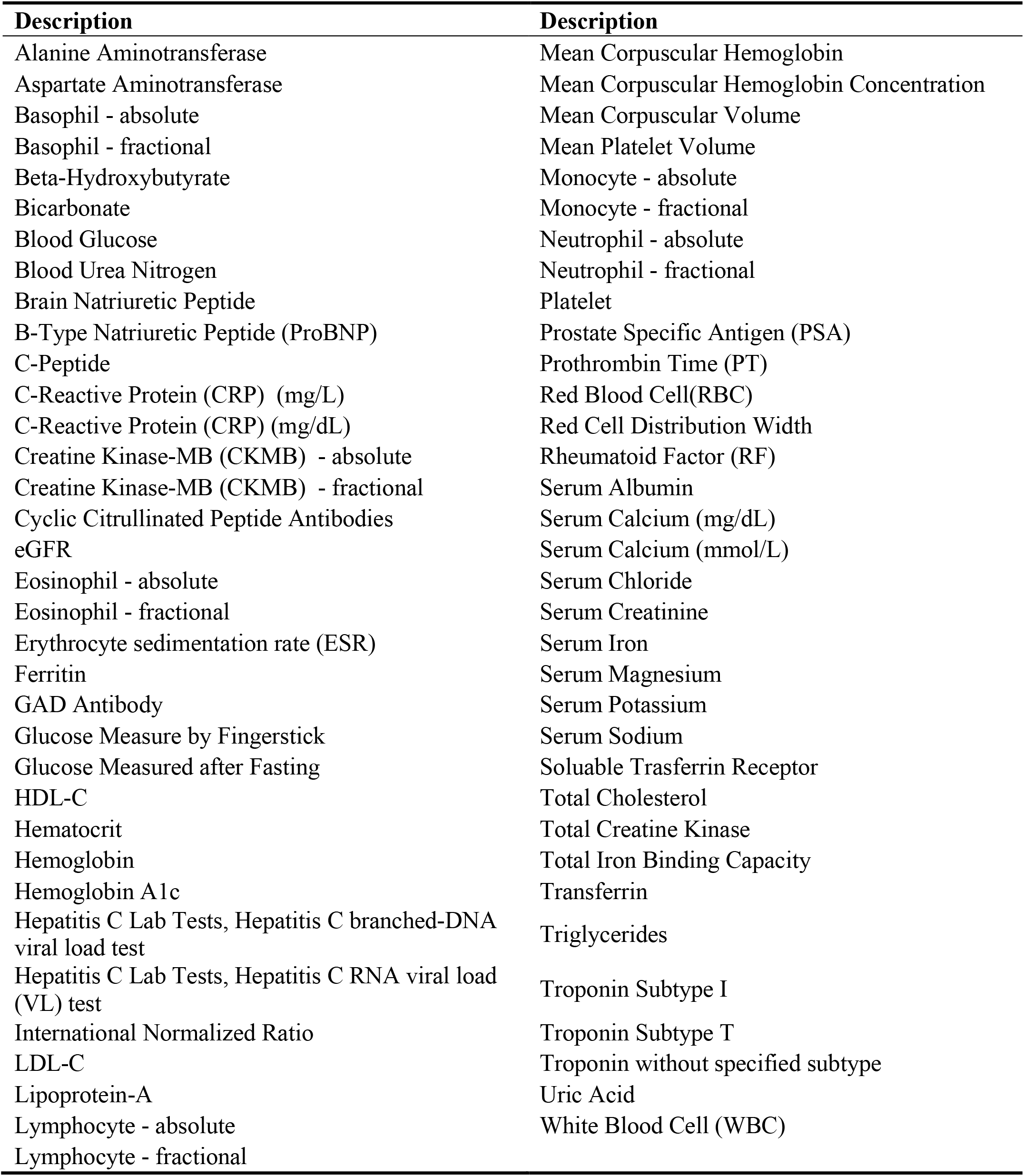
Curated laboratory measurements available in MVP (n=69).

| Description | Description |
| --- | --- |
| Alanine Aminotransferase | Mean Corpuscular Hemoglobin |
| Aspartate Aminotransferase | Mean Corpuscular Hemoglobin Concentration |
| Basophil - absolute | Mean Corpuscular Volume |
| Basophil - fractional | Mean Platelet Volume |
| Beta-Hydroxybutyrate | Monocyte - absolute |
| Bicarbonate | Monocyte - fractional |
| Blood Glucose | Neutrophil - absolute |
| Blood Urea Nitrogen | Neutrophil - fractional |
| Brain Natriuretic Peptide | Platelet |
| B-Type Natriuretic Peptide (ProBNP) | Prostate Specific Antigen (PSA) |
| C-Peptide | Prothrombin Time (PT) |
| C-Reactive Protein (CRP) (mg/L) | Red Blood Cell(RBC) |
| C-Reactive Protein (CRP) (mg/dL) | Red Cell Distribution Width |
| Creatine Kinase-MB (CKMB) - absolute | Rheumatoid Factor (RF) |
| Creatine Kinase-MB (CKMB) - fractional | Serum Albumin |
| Cyclic Citrullinated Peptide Antibodies | Serum Calcium (mg/dL) |
| eGFR | Serum Calcium (mmol/L) |
| Eosinophil - absolute | Serum Chloride |
| Eosinophil - fractional | Serum Creatinine |
| Erythrocyte sedimentation rate (ESR) | Serum Iron |
| Ferritin | Serum Magnesium |
| GAD Antibody | Serum Potassium |
| Glucose Measure by Fingerstick | Serum Sodium |
| Glucose Measured after Fasting | Soluble Transferrin Receptor |
| HDL-C | Total Cholesterol |
| Hematocrit | Total Creatine Kinase |
| Hemoglobin | Total Iron Binding Capacity |
| Hemoglobin A1c | Transferrin |
| Hepatitis C Lab Tests, Hepatitis C branched-DNA viral load test | Triglycerides |
| Hepatitis C Lab Tests, Hepatitis C RNA viral load (VL) test | Troponin Subtype I |
| International Normalized Ratio | Troponin Subtype T |
| LDL-C | Troponin without specified subtype |
| Lipoprotein-A | Uric Acid |
| Lymphocyte - absolute | White Blood Cell (WBC) |
| Lymphocyte - fractional |  |

## Statistical Methodology

### False discovery rate (FDR) controlled heterogeneity testing (hetFDR)

In this section, we present the implementation details of our proposed false discovery rate (FDR) controlled heterogeneity testing (hetFDR) approach.

### Notation and Setup

Let *J* be the number of ancestry groups, and *K* be the number of outcomes in association testing (phenotypes or laboratory values). For each subject *i* belonging to the ancestry group *j* ∈ {1,2, …, *J*} with *n*_*j*_ subjects, let 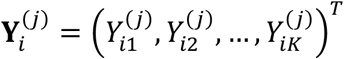 be the *K*-dimensional binary outcome vector of subject 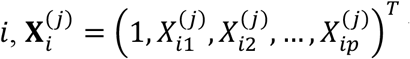 be the (*p* + 1)-dimensional adjustment covariates (e.g. age, and gender) vector including 1 for intercept, and 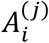 be the exposure variable (i.e. in this study the IL6R variant). Let

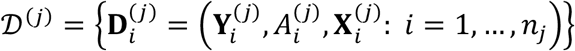

denote the data set of each ancestry group *j*. To characterize the association between 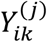 and 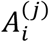, we introduce the logistic model:

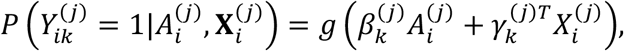

where *g*(*a*) = *e*^*a*^/(1 + *e*^*a*^) represents the logistic link function. For phenotypes *k* = 1,2, …, *K*, we aim at simultaneously testing for heterogeneity of the effect 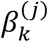 across the *J* ancestry groups:

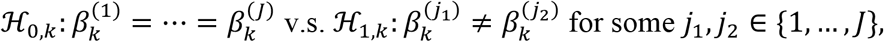

with the false discovery rate (FDR) controlled below some level *η* (e.g., *η* = 0.1):

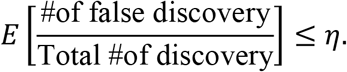

### Constructing Test Statistics

We first construct the effect estimator 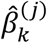 for each (*k, j*) and its asymptotic variance 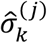 using the standard score test. Then for each phenotype *k*, we introduce a mean effect statistic constructed as the inverse-variance weighted average of 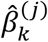 across ancestry groups:

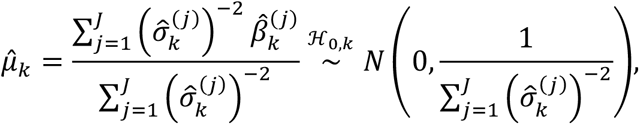

as well as a heterogeneity statistic constructed as the sample variance of 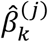 among ancestry groups:

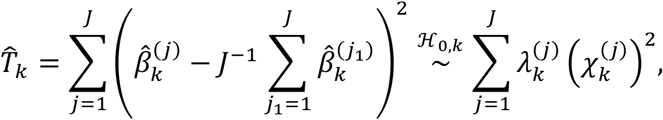

where 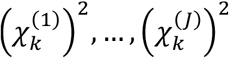 are independent chi-squared random variables with degree of freedom 1, and 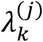’s are estimated by extracting the eigenvalues of the *J* × *J* empirical covariance matrix:

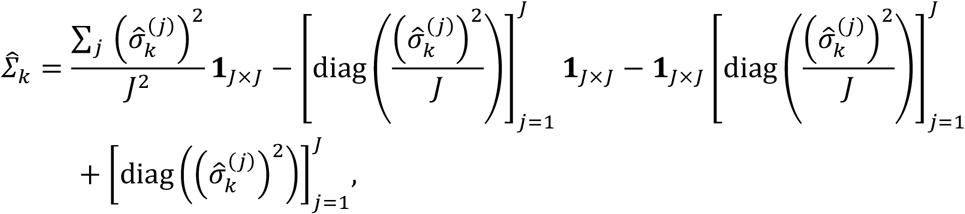

with **1**_*J*×*J*_ representing the *J* × *J* matrix of all ones. Here the heterogeneity test statistic 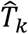 is actually a quadratic form of Gaussian random variables. Its corresponding *p*-value 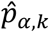 can be computed using the *CompQuadForm* package in **R** (de Micheaux, 2017). Our testing and multiple testing of heterogeneity is carried based on 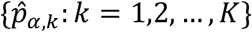. Meanwhile, we extract the *p*-value of the mean effect statistics 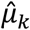, denoted as 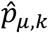, as a guiding information to improve the power of the multiple testing; see the next section for details. It is important to note that under our construction, 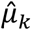 and 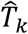 are asymptotically independent, which grants the validity of multiple testing with 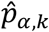 assisted by 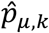.

### Weights Construction

We use 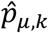 as a prior guidance to assign weights to 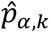, based upon the prior assumption that the non-null set of heterogeneity effects is close to that of the mean effects. Note that the effective sample size of 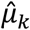 and 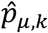 is the total sample size 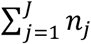 of all ancestry groups, while that of 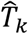 and 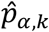 is actually dominated by the minority ancestry groups with small sample size. Thus, 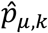 tends to provide a more precise information about the set of outcomes with non-zero mean effects, potentially serving as good side information to aid testing of the heterogeneity effects.

Inspired by recent literature of adaptive multiple testing that (Li and Barber, 2019; Cai et al., 2020, e.g.) leverages side guidance to enhance the power in comparison with the standard Benjamini Hochberg (BH) procedure (Benjamini and Hochberg, 1995), we propose the following procedures to convert 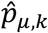 into proper weights of the candidate *p*-values 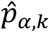:

1. Calculate 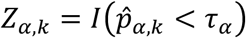 where *τ*_*α*_ is some pre-specified cutoff parameter. Practically, one can either fix *τ*_*α*_ as some small value like 10^−4^ or specify it empirically, e.g., choosing *τ*_*α*_ as the *p*-value cutoff returned from the BH procedure on 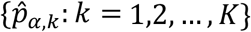 with level 0.5 (Cai et al., 2020).
2. Implement logistic regression on *Z*_*α,k*_ against 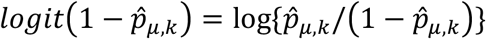 to obtain the intercept *a*_1_ and coefficient *a*_0_. And set

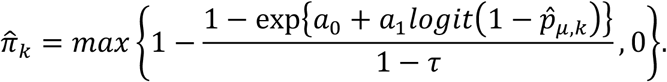
3. Standardize 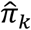 and obtain the final weights 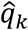 through: 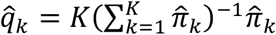.

Our construction of 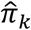 is motivated by the idea to find the non-null prior probability and the bayesian decision rule for each *k* as used in Cai et al. (2020). And our third step to standardize 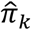 is used to protect the validity and FDR control.

### Adaptive FDR Control

Finally, we weight and adjust the heterogeneity testing *p*-values as 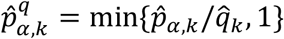 for *k* = 1,2, …, *K*, and implement the following algorithm for discovery with FDR control of level *η*.

#### Algorithm 1.

Adaptive multiple testing with FDR level *η*.

1. Find 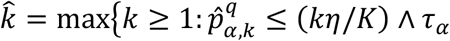 for at least *k* many 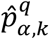};
2. Reject null hypothesis ℋ_*k*_ with 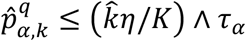 for a total of 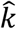 rejections.

### Simulation Studies

We design simulation studies to evaluate the proposed hetFDR approach and compare it with existing commonly used FDR control methods including Benjamini and Hochberg’s procedure (BHq) (Benjamini and Hochberg, 1995) and Stoery’s procedure (Storey, 2002). To mimic our real application, each simulated dataset consists of *K* = 1000 phenotypes, *J* = 2 ancestry groups, and *n*_*j*_ = 500 samples for each population. The set of non-null mean effects is set as *S*_*µ*_ = {1,2, …, 50}. Corresponding to our assumption, we set *S*_*α*_ as the first |*S*_*α*_| indices in *S*_*µ*_ so that *S*_*α*_ ⊆ *S*_*µ*_.

For each ancestry group *j* ∈ {1,2}, we generate the exposure *A*^(*j*)^ ∼ Bernoulli(0.5) and the covariates *X*^(*j*)^ ∼ *N*(0, *I*_*p*_) with *p* = 3. Then for the phenotypes *k* belonging to the null set {1,2, …, *K*}\*S*_*µ*_, we generate 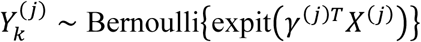 for *j* = 1,2, where *γ* = (0.25,0.25,0.25)^*T*^. For the phenotypes *k* ∈ *S*_*µ*_\*S*_*α*_, we generate 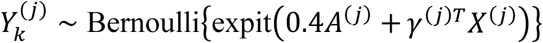 for *j* = 1,2. And for the phenotypes belonging to the heterogeneity set, i.e. *k* ∈ *S*_*α*_, we generate 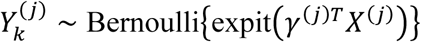 for *j* = 1 and 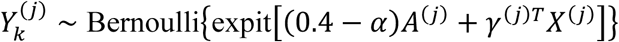 for *j* = 2. We implement two set of simulation settings, firstly, we fix *α* = 0.3 and let the number of heterogeneous effects |*S*_*α*_| vary in {10,25,40}. Second, we fix |*S*_*α*_| = 40 and let *α* vary in {0.2,0.3,0.4}. The desirable FDR level is set as 0.1 and the FDR and average power of all methods are estimated via 500 times of simulations in each setting.

The resulted FDR and average power are presented in the following Figure. Under different settings of the effect magnitude *α* and the number of heterogeneous effects |*S*_*α*_|, our proposed hetFDR method controls FDR below 0.1 and shows substantial and consistent higher average power than BHq and Storey’s procedures. For example, when |*S*_*α*_| = 25 and *α* = 0.3, hetFDR has about 0.3 higher power than the other two methods. This is because that our method additionally leverages the mean effect 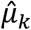 and its *p*-value as side information and assigns higher chances of rejection to the phenotypes belonging to *S*_*µ*_. Since *S*_*α*_ ⊆ *S*_*µ*_ and |*S*_*µ*_| is much smaller than *K*, this successfully reduces the price of screening out a large number of phenotypes with null effects.

One may also note that our method is conservative on FDR control when |*S*_*α*_| becomes larger. As an example, its FDR is around 0.05 when |*S*_*α*_| = 40, much smaller than the nominal level 0.1 achieved by the other methods. We believe this is due to that our weighting procedure can significantly restrict the candidate set to *S*_*µ*_ by largely down-weighting the *p*-values of phenotypes with large 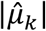. Then when *S*_*α*_ takes large proportion (80% when |*S*_*α*_| = 40) in *S*_*µ*_, our Algorithm 1[alg:1] will be conservative since it is a BHq-type procedure using the total number of hypotheses (effectively close to |*S*_*µ*_| in our method) to approximate the number of false discoveries.

**Figure:**
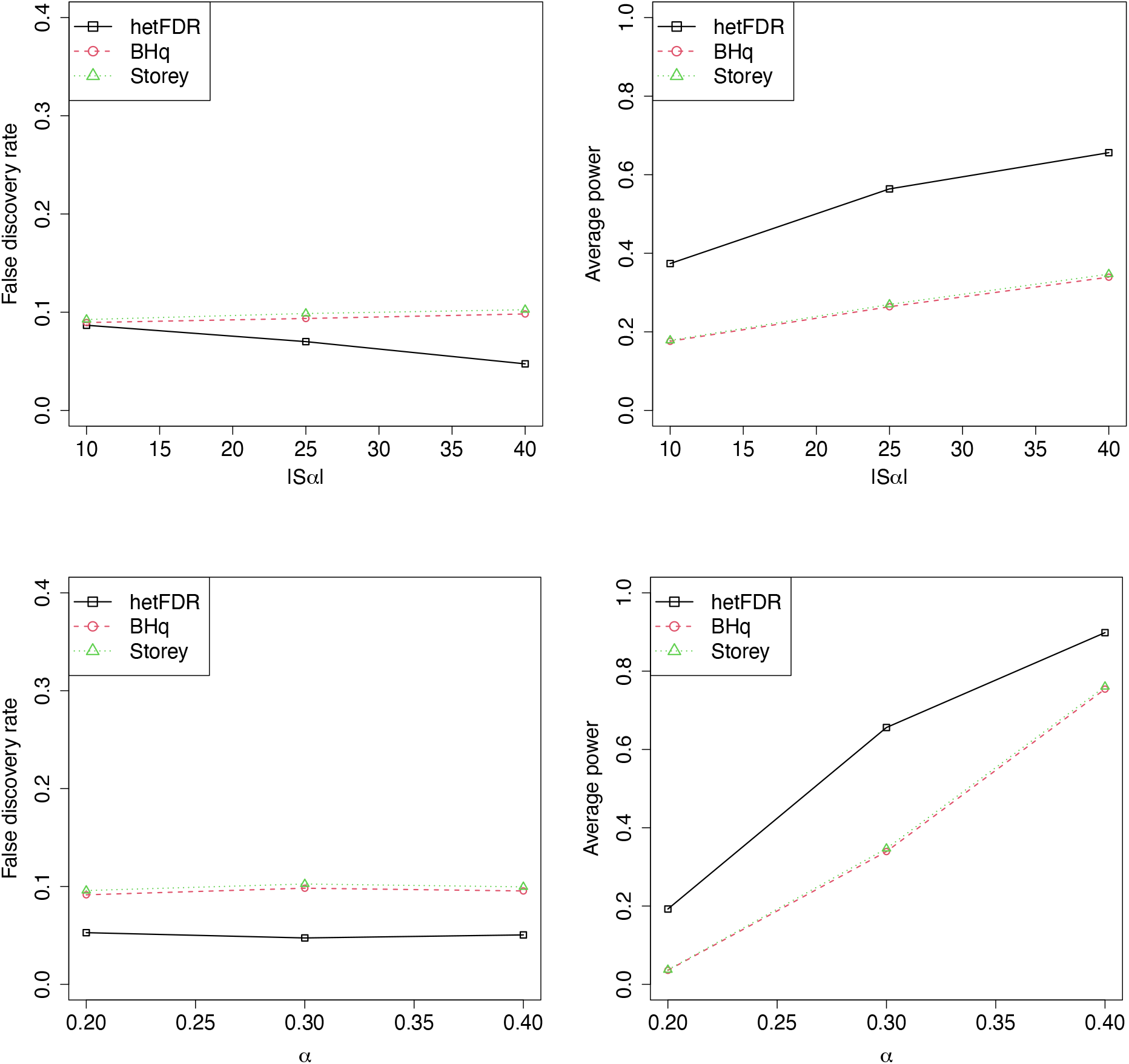
FDR and average power plotted against different choices of α or |Sα|. Methods under comparison include our proposed hetFDR, the standard BHq procedure, and Storey’s procedure. All values are estimated based on 500 times of simulations.

